# Forecasting the Cumulative Number of COVID-19 Deaths in China: a Boltzmann Function-based Modeling Study

**DOI:** 10.1101/2020.03.02.20030064

**Authors:** Yuanyuan Gao, Zhongyan Li, Qi Ying, Cheng Long, Xinmiao Fu

**Author notes:** To whom correspondence should be addressed to Professor Xinmiao Fu and Dr. Cheng Long. These authors contributed to this work equally (Yuanyuan Gao, Zhongyan Li).

## Abstract

The COVID-19 outbreak is on-going in China. Here we estimated the potential total numbers of COVID-19 deaths in China, outside Hubei (in China), Hubei Province, Wuhan City and outside Wuhan (in Hubei) by Boltzmann function-based analyses, which are 3342 (95% CI, 3214, 3527), 111 (109, 114), 3245 (3100, 3423), 2613 (2498, 2767) and 627 (603, 654), respectively. The results may help to evaluate the severity of COVID-19 outbreaks and facilitate timely mental service for the families of passed patients.

---

An outbreak of 2019 novel coronavirus diseases (COVID-19) caused by SARS-CoV-2 is on-going in China and has spread worldwide ^[1, 2]^. As of Feb 13, 2020, there have been over 80000 confirmed COVID-19 patients and over 3000 deaths in China, most of which are in the epicenter of the outbreak, Wuhan city and related regions in Hubei province. Although the number of new confirmed cases has substantially decreased since Feb 13, 2020 and the outbreak appears to approach the late phase in China, people have raised grave concerns about the severity of the outbreak, especially questioning how many patients will die eventually. Here we estimated the potential total number of COVID-19 deaths by applying Boltzmann function-based regression analysis, an approach we recently developed for estimating the potential total numbers of confirmed cases for both the ongoing SARS-CoV-2 outbreak and the gone 2003 SARS epidemic ^[3]^.

We collected data for analysis on the officially released cumulative numbers of deaths in mainland China, other provinces than Hubei, Hubei Province, Wuhan City, and other cities in Hubei (from Jan 21 to Feb 29, 2020). We first verified that the cumulative numbers of confirmed cases with respect to each region were all well fitted to the Boltzmann function (*R*^*2*^ all being close to 0.999); **Fig. 1A**), consistent with our earlier report using the data from Jan 21 to Feb 14, 2020 ^[3]^. Assuming that the number of deaths is proportional to the number of confirmed cases for the outbreak under specific circumstances, we speculated that the cumulative number of COVID-19 deaths would also obey the Boltzmann function. In support of this speculation, the cumulative numbers of COVID-19 deaths in the above regions were all well-fitted to the Boltzmann function (*R*^*2*^ all being close to 0.999; **Figs. 1B, 1C** and **Table 1**), with the potential total numbers of deaths being estimated as 3200±40, 108±1, 3100±40, 2500±40 and 604±6 respectively (**Table 1**). This result, in conjunction with our earlier observation that the cumulative numbers of confirmed cases of 2003 SARS in mainland China and worldwide were well fitted with the Boltzmann function, prompted us to analyze the cumulative numbers of 2003 SARS deaths in the same way. Consistently, we observed that the cumulative numbers of 2003 SARS deaths in mainland China, Hong Kong and worldwide were all well fitted to the Boltzmann function (**Fig. 1D**), strongly suggesting that the Boltzmann function is suitable to simulate the course of deaths associated with coronavirus-caused diseases.

**Table 1.** Summary of the estimated total numbers of COVID-19 deaths in China

| Regions | Boltzmann <sup>a</sup> |  | Boltzmann <sup>a</sup><br>(mean, 95% CI) | Richards <sup>a</sup><br>(mean, 95% CI) |
| --- | --- | --- | --- | --- |
| | Mean±SD | $R^2$ | | |
| mainland China | 3200±40 | 0.999 | 3260 (3187, 3394) | 3342 (3214, 3527) |
| other provinces | 108±1 | 0.996 | 110 (109, 112) | 111 (109, 114) |
| Hubei Province | 3100±40 | 0.999 | 3174 (3095, 3270) | 3245 (3100, 3423) |
| Wuhan City | 2490±40 | 0.998 | 2550 (2494, 2621) | 2613 (2498, 2767) |
| other cities in Hubei | 604±6 | 0.999 | 617 (607, 632) | 627 (603, 654) |

<sup>a</sup> Boltzmann function-based regression analysis results assuming the reported cumulative number of deaths are precise and have uncertainty (a standard deviation of 2.5%), respectively (for detail, refer to the Methods section in the supporting information file and **Figs. 1E, S1**). The same uncertainty was set for Richards function-based regression analysis (for detail, refer to the Methods section and **Fig. S2**)

**Figure 1.**
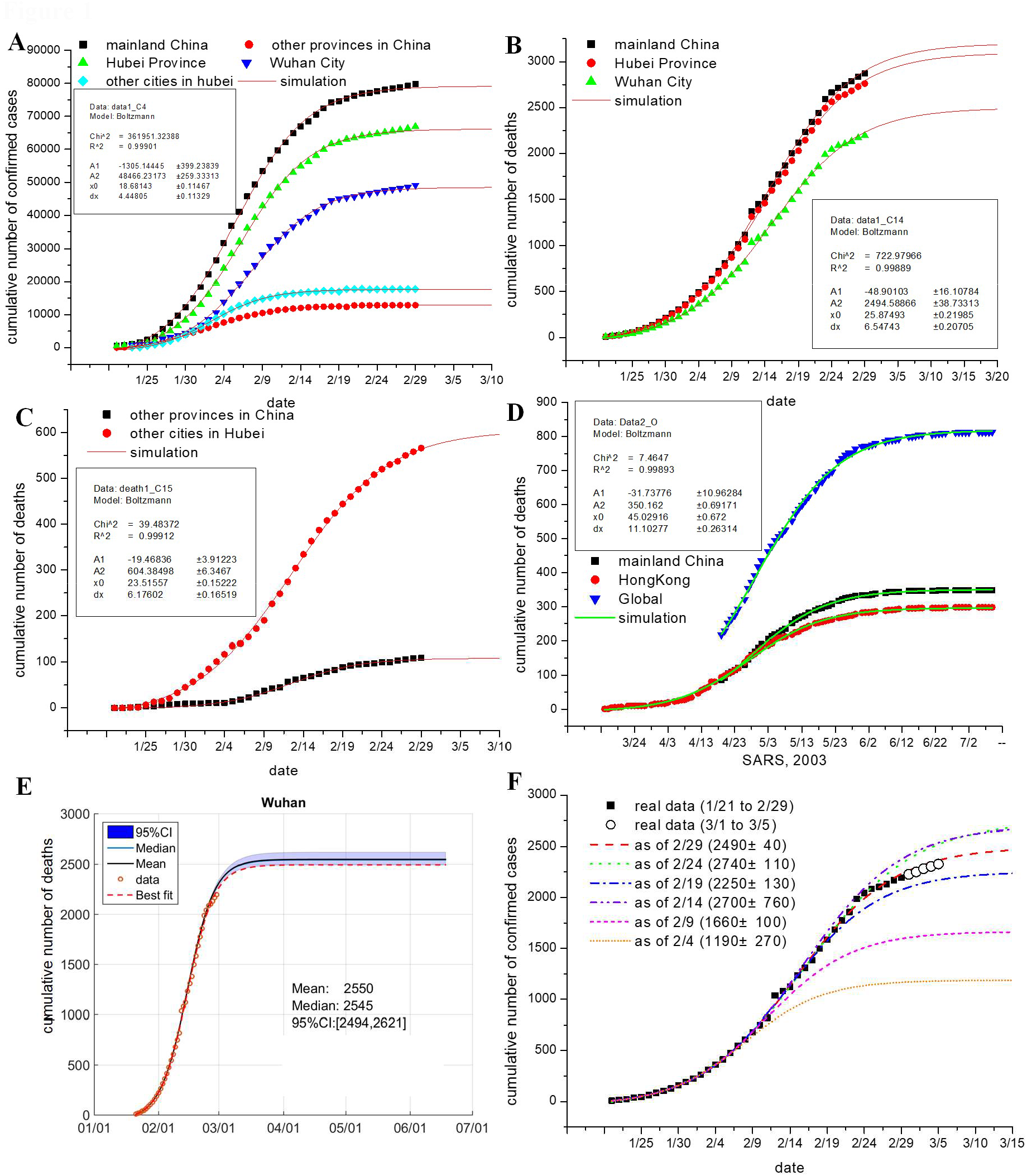
Fitting the cumulative number of COVID-19 deaths to Boltzmann function. (**A, B, C**) Boltzmann function-based regressions analysis results on the cumulative numbers of confirmed COVID-19 cases (panel A) and deaths (panels B and C) in the indicated geographic regions. Parameters of the established functions for Wuhan City (panels A and B) and for other cities in Hubei (panel C) are shown in insets. Note: the reported cumulative number of confirmed cases of Hubei Province and Wuhan City were re-adjusted for data fitting due to the suddenly added cases determined using clinical features (for details, refer to **Table S1**). (**D**) Boltzmann function-based analysis results on the cumulative numbers of 2003 SARS deaths in the indicated regions. Parameters of the established function for mainland China are shown in insets. (**E**). Regressions analysis results on COVID-19 deaths in Wuhan City by the Boltzmann functions assuming that the relative uncertainty of the data follows a single-sided normal distribution with a mean of 1.0 and a standard deviation of 2.5%. Original data are shown as circles; simulated results are presented as colored lines as indicated. Inserts show key statistics. Results for other regions are presented in **Figs. S1**. (**F**). Prediction of COVID deaths in Wuhan City by Boltzmann function-based analyses. The real data from Jan 21 to different ending dates were arbitrarily analyzed (colored lines), with the potential total numbers of deaths under these analyses being shown in insets.

One issue regarding our analyses is that some COVDI-19 deaths might be miss-reported such that the reported death numbers represent a lower limit. For instance, 134 new deaths were suddenly counted from more than 13000 clinically diagnosed patients in Hubei Province on Feb 12, 2020 (as clearly indicated by a sudden jump of deaths in **Fig. 1B**). Another uncertainty might result from those unidentified COVID-19 deaths at the early phase of the outbreak. We applied the Monte Carlo method (for detail, refer to the Methods section in SI file) to estimate such uncertainty assuming that the relative uncertainty of the reported numbers of deaths follows a single-sided normal distribution with a mean of 1.0 and a standard deviation of 2.5%. The potential total numbers of COVID-19 deaths in the above regions were estimated to be 3260 (95% CI 3187, 3394), 110 (109, 112), 3174 (3095, 3270), 2550 (2494, 2621) and 617 (607, 632), respectively (**Figs. 1E** and **S1**), which are slightly higher than those estimated without uncertainty (refer to **Table 1**).

To verify our Boltzmann function-based estimations, we calculated the potential total numbers of deaths in the above regions by applying Richards function-based regression analyses, which had been explored to simulate the cumulative numbers of confirmed cases of 2003 SARS in different regions ^[4, 5]^. The potential total numbers of COVID-19 deaths in mainland China, other provinces, Hubei Province, Wuhan City and other cities were estimated to be 3342 (3214, 3527), 111 (109, 114), 3245 (3100, 3423), 2613 (2498, 2767) and 627 (603, 654), respectively (**Fig. S2**), which are close to what are estimated by the Boltzmann function-based analyses (**Table 1**). In addition, we found that the established Boltzmann function was able to predict the death course in a short period such that the released cumulative numbers of deaths from Mar 1 to 5 Mar 5 are close to the estimated numbers, as exemplified in Wuhan (**Fig. 1F** and **Table S1**). If the data from Jan 21 to different closing dates were arbitrarily analyzed, we found that the course of COVID-19 deaths could be largely simulated based on the data as of Feb 14 (**Fig. 1F**), implicating that the Boltzmann function-based analysis could predict the trend ahead of approximate three weeks.

Collectively, we observed that all sets of data from both the COVID-19 deaths and the 2003 SARS deaths were well fitted to the Boltzmann function. We propose that the Boltzmann function is suitable for analyzing not only the cumulative number of confirmed COVID-19 cases, as reported by us recently ^[3]^ (also refer to **Fig. 1A**), but also those of deaths as reported here. We noticed that modeling studies on the COVID-19 outbreak have been performed ^[6]^ and COVID-19 deaths have been estimated by other groups using different models. For instance, Li et al recently showed ^[7]^ by data driven analysis that a total of deaths in Hubei would be 2250, a number much lower than the observed (2761 as of Feb 29). Using the Susceptible-Infected-Recovered-Dead model Anastassopoulou et al forecasted that the total death might exceed 7000 by Feb 29 ^[8]^, a number apparently much higher.

Since case fatality ratio in the epicenter of the outbreak is still much higher than that in other provinces of mainland China ^[1, 2]^, there is a great potential for government to optimize preparedness and medical resource supplies therein, by which hundreds of lives of COVID-19 patients, particularly those severe and critically ill patients ^[2, 9]^, might be saved. In addition, our estimates on the course of COVID-19 deaths (refer to **Table S2**) may benefit the mental health service that needs to be timely provided to the families of passed patients ^[10]^.

## Data Availability

all data are included in the manuscript and supplementary data file.

## Acknowledgments

This work is support by the National Natural Science Foundation of China (No. 31972918 and 31770830 to XF).

